# HIV prevalence in transgender women and cisgender men who have sex with men in sub-Saharan Africa 2010-2022: a meta-analysis

**DOI:** 10.1101/2023.11.09.23298289

**Authors:** Oliver Stevens, Rebecca L. Anderson, Keith Sabin, Sonia Arias Garcia, Elizabeth Fearon, Kingsley Manda, Wame Dikobe, Lloyd Mulenga, Neena M. Philip, Mathieu Maheu-Giroux, Jinkou Zhao, Mary Mahy, Jeffrey W. Imai-Eaton

**Affiliations:** MRC Centre for Global Infectious Disease Analysis, School of Public Health, Imperial College London, London, United Kingdom; Data for Impact, The Joint United Nations Program on HIV/AIDS (UNAIDS), Geneva, Switzerland; University College London, London, United Kingdom; Malawi National Statistics Office, Zomba, Malawi; FHI360, Gaborone, Botswana; Zambia Ministry of Health, Lusaka, Zambia; ICAP, Columbia University Mailman School of Public Health, New York, New York, USA; Department of Epidemiology and Biostatistics, School of Population and Global Health, McGill University, Montréal, Canada; The Global Fund to Fight AIDS, Tuberculosis and Malaria, Geneva, Switzerland; Center for Communicable Disease Dynamics, Department of Epidemiology, Harvard T.H. Chan School of Public Health, Boston, MA, USA

**Author notes:** Corresponding author: Oliver Stevens, St. Mary’s Hospital Campus Norfolk Place London W2 1PG United Kingdom.

**Keywords:** HIV-1, Transgender persons, Homosexuality, Male, Sexual and Gender Minorities, Africa

## Abstract

**Introduction:** The Global AIDS Strategy 2021-2026 calls for equitable access to HIV services for all populations. Transgender people have been marginalised and experience disproportionate risk of HIV infection in sub-Saharan Africa (SSA) and data to guide HIV programmes are severely limited. Surveillance data among cisgender men who have sex with men (cis-MSM) are comparatively abundant. We assessed whether HIV prevalence among cis-MSM was correlated with HIV prevalence among transgender women.

**Methods:** Data from key population surveys conducted in SSA between 2010-2022 were identified from existing databases and survey reports. Studies that collected HIV prevalence on both transgender women and cis-MSM populations were analysed with random effect meta-analysis to estimate the ratio of HIV prevalence among cis-MSM:transgender women.

**Results:** Twenty-one studies were identified encompassing 8,476 transgender women and 24,102 cis-MSM. Median HIV prevalence among transgender women was 23.5% (interquartile range [IQR] 11.5-39.8%) and 16.2% (IQR 8.1-26.8%) among cis-MSM. HIV prevalence among transgender women was 50% higher than in cis-MSM (prevalence ratio 1.48 95CI 1.25-1.76). HIV prevalence among transgender women was highly correlated with year/province-matched HIV prevalence among cis-MSM (R^2^=0.60), but poorly correlated with year/province-matched total population HIV prevalence (R^2^=0.01).

**Conclusion:** Transgender women experience a significantly greater HIV burden than cis-MSM in SSA, underscoring the need for HIV services addressing the disproportionate vulnerability experienced by transgender women. Further bio-behavioural surveys focused on determinants of HIV infection, treatment uptake, and risk behaviours among transgender people, distinct from cis-MSM, will improve understanding of HIV risk and vulnerabilities.

## Introduction

The Global AIDS Strategy 2021-2026 calls for equitable and equal access to HIV prevention and treatment services for all to reduce HIV incidence and end HIV/AIDS as a public health threat by 2030^1^. The Strategy has motivated renewed focus to quantify population size, HIV prevalence, ART coverage, and new HIV infections among key populations in sub-Saharan Africa (SSA) to reduce inequality in the global HIV/AIDS response.

Transgender people experience a HIV risk environment wherein the combination of exposure to efficient HIV transmission routes and socio-structural risk factors result in disproportionate HIV burden^2–5^. Societal marginalisation, criminalisation, and stigma have impeded collection of robust HIV surveillance data among key populations in sub-Saharan Africa^6–8^. Global systematic reviews of HIV epidemiology among transgender people disproportionately reflect data outside SSA^4,5^. Only eleven SSA countries have population size estimates for transgender women, twenty countries have surveillance data on HIV prevalence, and nine have ART coverage data^9^.

HIV prevalence data for men who have sex with men (MSM) in SSA are more widely available. Over 350 HIV prevalence estimates have been measured since 2010 across 32 of 38 SSA countries^9^. While cis-MSM and transgender women have distinct vulnerabilities to HIV^10,11^, studies in SSA indicate that HIV epidemiology in these two populations is linked: transgender women are recruited by cis-MSM into surveys using social and sexual network recruitment^12,13^; transgender women most often report male sexual partners and receptive anal sex^12,14,15^; phylogenetic analysis indicates HIV transmission clustering between transgender women and cis-MSM^16,17^; and qualitative data indicate that some transgender people identify as MSM^18,19^. Many surveillance studies primarily collecting data among cis-MSM have also included transgender people, providing a source of data in the absence of studies explicitly aiming to recruit transgender people.^20,21^ We sought to meta analyse relative HIV prevalence between cis-MSM and transgender women. This may be used to highlight the high HIV burden among transgender women in settings that lack surveillance data among this key population.

## Methods

We extracted data on HIV prevalence among transgender people and cis-MSM between 2010-2022 in sub-Saharan Africa from UNAIDS Global AIDS Monitoring^22^, The Global Fund surveillance database, US Centers for Disease Control and Prevention key population surveillance databases, and literature searches^9^. Studies that reported estimates of HIV prevalence for cis-MSM and any transgender population (as defined by study authors; Supplementary Table S1) in the same study were considered for inclusion. Tabulations of relevant data not reported in study publications were shared by the authors of Fearon et al.^13^, Keshinro et al.^24^, and Malawi 2019-20, Botswana 2017, and Zambia 2021 Biobehavioural Surveys^25–27^.

HIV prevalence data were pooled and weighted proportional to the number of individuals tested. Where studies reported HIV prevalence for more than one transgender population, HIV prevalence among transgender women only was included in the meta-analysis. Log-normal random-effects meta-analysis was used to calculate pooled prevalence ratios (PRs) for HIV prevalence among transgender women compared to cis-MSM. Estimates were calculated separately for Eastern and Southern Africa (ESA) and Western and Central Africa (WCA) regions, and across all sub-Saharan Africa using nested study, country, and area random effects. We conducted a sensitivity analysis that included a fixed effect for calendar year, centred at the mean year of study. Analyses were conducted in R 4.1.2^28^, using the *metafor* package^29^.

Province-level estimates of sex-specific total population HIV prevalence were extracted from UNAIDS estimates and projected backward 2000-2020 parallel to national-level HIV prevalence trajectories.^30,31^ Four additional observations of HIV prevalence among transgender women^14,35–37^ were included in in correlation analysis with total population HIV prevalence, which had been excluded from cis-MSM prevalence ratios meta-analysis because they lacked matching cis-MSM HIV prevalence estimates.

This study received ethical approval from the Imperial College Research and Ethics Committee (#6473559). Data are available at https://doi.org/10.5281/zenodo.10838437

## Results

Twenty-one studies (11 in ESA and 10 in WCA) were included in the analysis, encompassing 8,476 transgender women and 24,102 cis-MSM (Supplementary Table S1). Eleven studies were from journal publications and ten were identified from grey literature including biobehavioural survey (BBS)^38^ and PLACE study^39^ reports. The median denominator for HIV prevalence among transgender women was 89 (IQR 35-235) and 368 (IQR 198-426) for cis-MSM. HIV status was ascertained by serological assays in all studies. Median HIV prevalence among transgender women in ESA was 25.0% (IQR 10.5-42.2%) and 23.3% (IQR 14.1-35.2%) in WCA. Median HIV prevalence among cis-MSM in ESA was 15.2% (IQR 8.8-24.6%) and 17.0% (IQR 7.8-27.5%) in WCA.

Pooled across sub-Saharan Africa, HIV prevalence in transgender women was 1.48 times higher (95%CI 1.25-1.76) than in cis-MSM (Figure 1). In ESA, prevalence among transgender women was 1.49 times higher than cis-MSM (95%CI 1.26-1.75) and in WCA 1.38 times higher (95%CI 0.92-2.07). In sensitivity analysis, HIV prevalence ratio declined 6% per year, but this was not statistically significant (annual PR 0.94; 95%CI 0.89-1.00).

**Figure 1:**
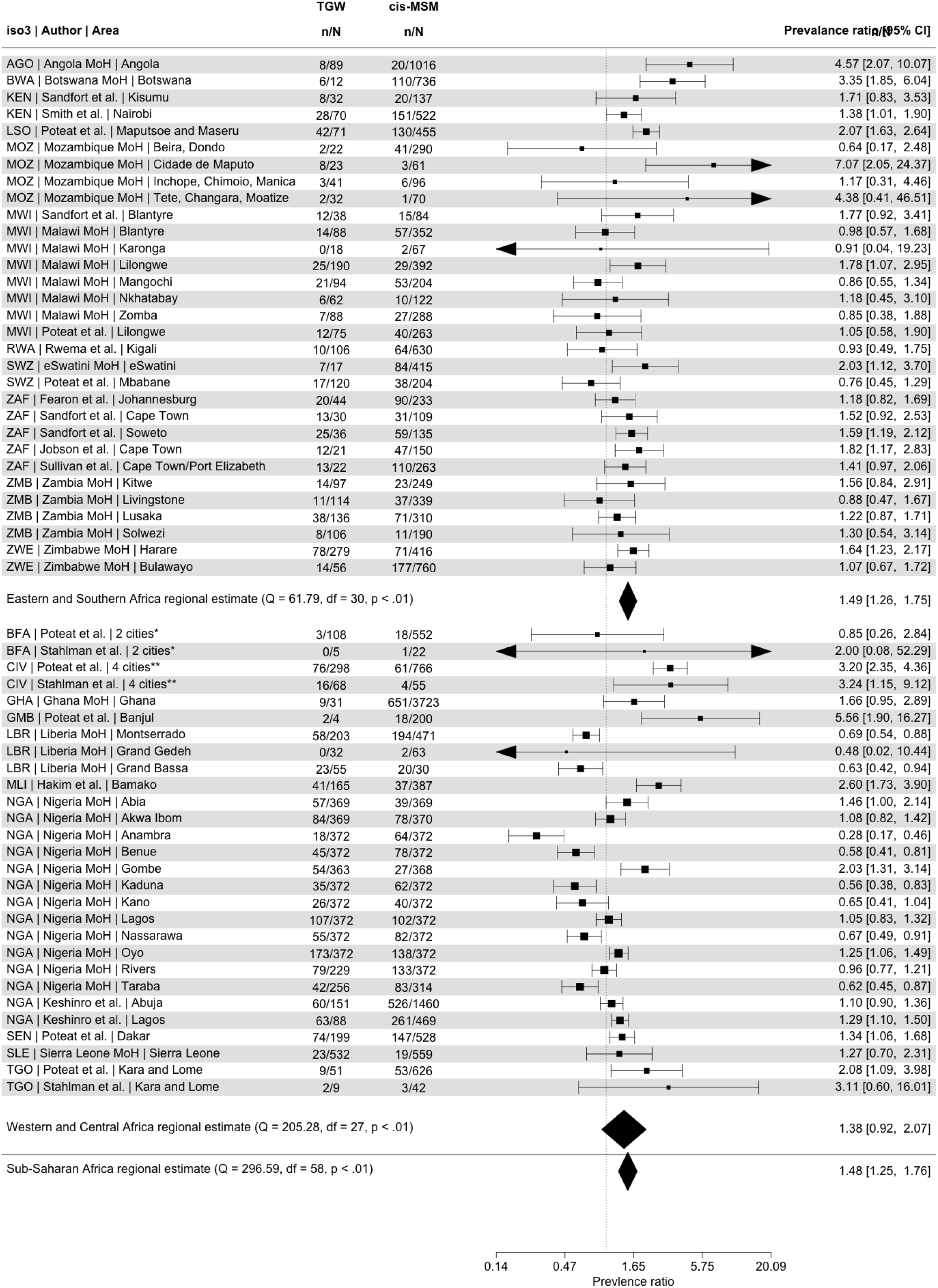
Forest plot comparing HIV prevalence ratios in transgender women (TGW) and cis-gendered men-who-have-sex-with-men (cis-MSM). * Bobo-Dioulasso and Ouagadougou ** Abidjan, Bouake, Gagnoa, and Yamoussoukro

HIV prevalence among transgender women was highly correlated with study-matched cis-MSM HIV prevalence (Figure 2; R^2^=0.60) and poorly correlated with province/year-matched total population HIV prevalence (R^2^=0.01). Relative to venue- and convenience-based sampling methods, studies using network-sampling methods did not estimate a significantly different pooled HIV prevalence ratio (Supplementary Table S1; PR 1.06 95%CI 0.72-1.56).

**Figure 2:**
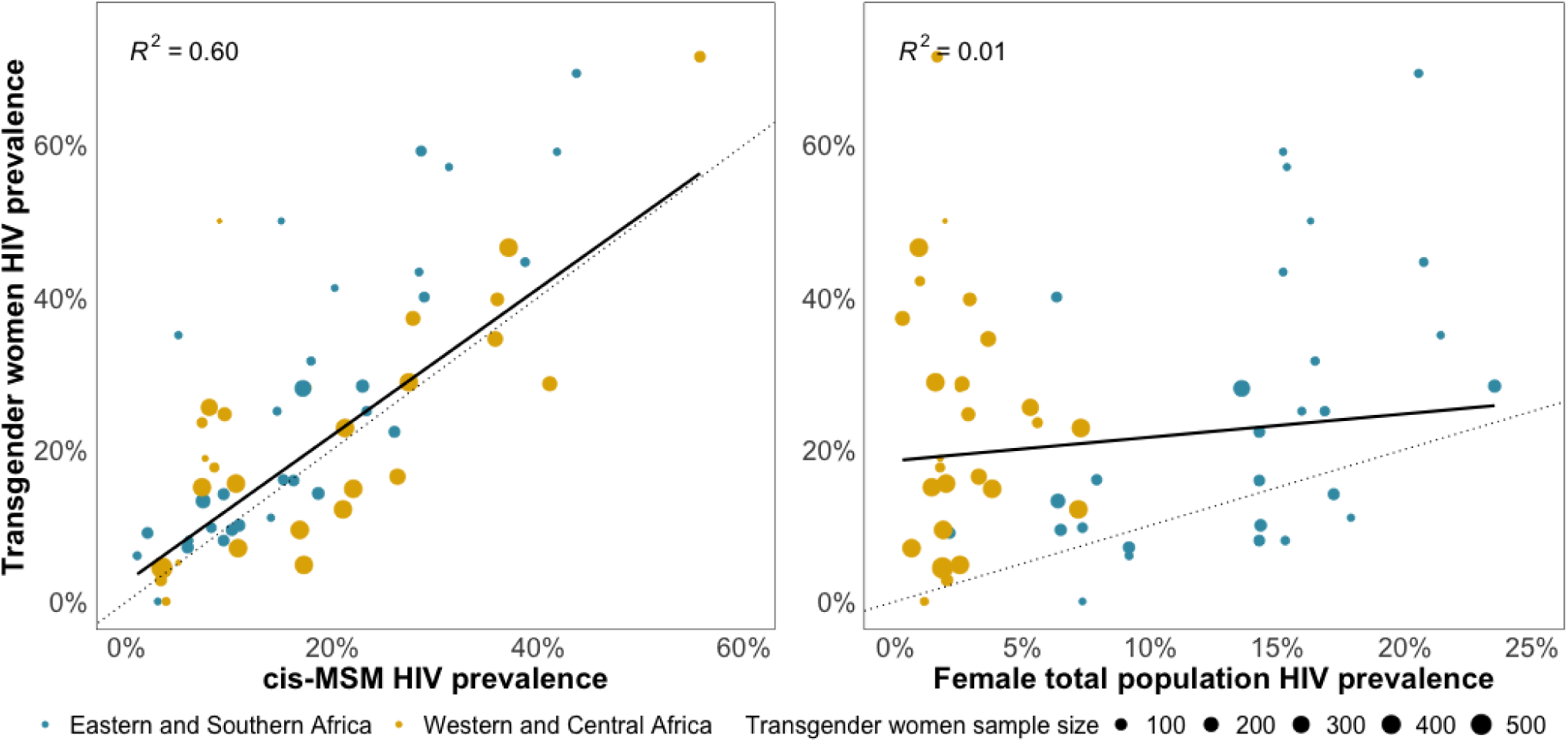
Comparison of HIV prevalence among transgender women and cis-gender men-who-have-sex-with men (cis-MSM) (left) and HIV prevalence among transgender women compared to HIV prevalence among all women aged 15-49 years (right). The dotted line represents the line of equality is (y=x). Colour of data points represents the UNAIDS region of the study country.

One-third of studies (6/21) included, or did not specifically exclude, transgender men in the sampled transgender population and were excluded from the primary meta-analysis. In sensitivity analysis excluding studies that reported combined HIV prevalence for transgender women and men, the estimated prevalence ratio was slightly higher (SSA PR: 1.63, 95%CI 1.36-1.96).

## Discussion

We identified twenty-five studies that measured HIV prevalence among transgender women, a large increase from ten African studies included in a 2022 systematic review^40^. HIV prevalence was 50% higher among transgender women than cis-MSM in sub-Saharan Africa, providing further evidence of HIV vulnerability distinct from cis-MSM. Previous studies^4,5,41^ have identified substantial HIV burden among transgender women, which this meta-analysis supports both in absolute terms and relative to cis-MSM. HIV service needs for transgender women are increasingly recognised in HIV programmes in sub-Saharan Africa and are identified in the National Strategic HIV/AIDS Plans of sixteen countries^42^, increasing from ten in 2018^43^. Our updated analysis included 21 studies compared to 8 studies identified in a 2017 review by Poteat et al.^20^ (a more recent 2023 review by Kloek et al.^41^ identified 10 studies among TGW but did not compare HIV prevalence with that among cis-MSM). Among those studies available in 2017, the prevalence ratio was 1.75 (95%CI 1.54-2.00), compared to 1.48 in this analysis.

We did not report pooled estimates of HIV prevalence among transgender women or relative risk of HIV infection compared to the total population^4,5^. Firstly, the relative risk of HIV infection in key populations varies with total population HIV prevalence in SSA. Summarising a single relative risk of infection decouples the key population epidemiology in a setting from the HIV epidemic in the total population. In WCA, cis-MSM HIV prevalence can be 100-fold higher than male total population prevalence, while in ESA it may be similar to, or lower than, that in the total population^9^. Second, total population HIV prevalence is poorly correlated with cis-MSM HIV prevalence, particularly in WCA where neighbouring countries with similar generalised HIV epidemics can have disparate key population epidemics^9,44^. Instead, we found a consistently higher risk of HIV infection in transgender women compared to cis-MSM. These estimates should be viewed as interim estimates to support programme planning in settings that lack dedicated HIV surveillance, and should not obviate the need for transgender-specific surveillance nor to further discourse that the programmatic needs of transgender people can be wholly met through cis-MSM programmes.^45^ Recognising the additional HIV burden experienced by transgender women and their specific health needs (e.g. gender-affirming hormone therapy), while identifying common needs among sexual and gender minorities (e.g. stigma and gender-based violence interventions) is an important strategy to ensure effective services reach those who need them in an era of constrained resources for key population programmes.

We were unable to estimate the relative HIV prevalence among transgender men relative to other populations, though limited data indicated that transgender men may have a lower prevalence of HIV infection than transgender women or cis-MSM (Supplementary Table S2)^16,51^ Of six studies in this analysis that did not exclude transgender men, four included recent oral or anal sex with a man as an inclusion criterion. Though data across SSA are limited, they indicate that cis-women are more likely to be partners of transgender men than cis-MSM or other transgender men^26,46,47^. Transgender men are likely, therefore, to be excluded from biological surveillance if all transgender populations are associated with the same HIV risk behaviours as cis-MSM.

We retained three studies that did not specify which transgender population(s) were recruited in the main analysis^25,48,49^. Sexual behaviour data collected in each study indicated that transgender participants overwhelmingly reported male sexual partners and receptive anal intercourse. All three studies were recent, published since 2020, and with large sample sizes. Sensitivity analysis indicates that their inclusion had only a marginal impact on the estimated relative risk of HIV infection, and we feel that their inclusion alongside studies that explicitly recruited transgender women is justified. Survey data on gender identity were often ambiguous, necessitating its removal from analysis further reducing sample sizes. Further research into accurately recording gender identity across diverse settings^19^ would improve gender identification in key population surveys and enhance comparability across studies^46,50^.

Few studies have measured treatment coverage or viral load suppression among transgender people, and only a subset is laboratory confirmed^9^. While we identified that transgender women are at increased risk of HIV infection, it was not possible to estimate ART coverage or access to treatment services either in absolute terms or relative to cis-MSM. This remains a key data gap in monitoring the objectives laid out by the Global AIDS Strategy.

Interpretation of this analysis is subject to several limitations. Firstly, we did not differentiate between studies whose primary objective was transgender surveillance^12,15,24,27,49,51–58^, and those that reported transgender participants alongside the primary objective of cis-MSM surveillance^13,25,26,48,59–61^, which may disproportionately recruit transgender people who socialise with cis-MSM. These data may overstate the similarities between the two populations as phylogenetic clustering studies indicate extensive transmission between cis-MSM/TGW and the heterosexual-identifying population^62^. Second, insufficient age-specific HIV prevalence data were available to estimate age-adjusted prevalence risk ratios which may lead to underestimated prevalence ratios.

## Conclusion

Transgender women have 50% higher HIV prevalence than cis-MSM in SSA and, therefore, transgender women and cis-MSM should not be considered as a single risk population. HIV prevalence among transgender women is highly correlated with cis-MSM prevalence, and cis-MSM HIV prevalence may help inform assessment of HIV prevalence among transgender women in settings that lack dedicated surveillance. Studies estimating the treatment cascade among transgender women and men are a priority to quantify progress towards equitable HIV service access and enable HIV programmes to meet the prevention and treatment needs of transgender people.

## Data Availability

All data produced in the present study are contained in the manuscript and at https://doi.org/10.5281/zenodo.10838437

## Competing interests

None

## Authors’ contributions

OS, KS, MM, and JWI-E conceptualised the study. KS, SAG, and JZ curated key population survey databases. EF, TC, KM, WD, and LM provided additional unpublished data from surveillance studies. OS and RA reviewed primary source documents, deduplicated data, and extracted data. OS analysed the data and wrote the first draft of the manuscript. All authors contributed to interpretation of results and edited the manuscript for intellectual content. All authors read and approved the final version of the manuscript for submission.

## Acknowledgments and funding

This research was supported by UNAIDS, the Bill and Melinda Gates Foundation (OPP1190661), National Institute of Allergy and Infectious Disease of the National Institutes of Health under award number R01-AI136664, and the MRC Centre for Global Infectious Disease Analysis (reference MR/R015600/1), jointly funded by the UK Medical Research Council (MRC) and the UK Foreign, Commonwealth & Development Office (FCDO), under the MRC/FCDO Concordat agreement and is also part of the EDCTP2 programme supported by the European Union.

---

**Supplementary Table S1:** Studies included in meta-analysis of HIV prevalence in transgender women (TGW) and cis-gendered men who have sex with men (cis-MSM)

| Study | Year of publication | Country | Year of data collection | Recruitment method | Transgender women inclusion criterion | TGW prevalence (%; n/N) | cis-MSM prevalence (%; n/N) |
| --- | --- | --- | --- | --- | --- | --- | --- |
| **Stahlman et al.**<sup>15</sup> | 2016 | Burkina Faso | 2013 | RDS | Identified as female or transgender, assigned male sex at birth, and reported having had insertive or receptive anal sex with a man in the past 12 months | 0.0% (0/5) | 4.5% (1/22) |
|  |  | Côte d'Ivoire | 2015 |  |  | 23.5% (16/68) | 7.3% (4/55) |
|  |  | Togo | 2013 |  |  | 22.2% (2/9) | 7.1% (3/42) |
| **Keshinro et al.**<sup>24</sup> | 2016 | Nigeria | 2016 | RDS | Identified as female, assigned male sex at birth, and reported having had insertive or receptive anal sex with a man in the past 12 months | 51.5% (123/239) | 40.7% (786/1929) |
| **Botswana MoH**<sup>26</sup> | 2017 | Botswana | 2017 | RDS | Identified as a woman, formerly or currently biologically male | 50.0% (6/12) | 14.9% (110/736) |
| **Ghana MoH**<sup>61</sup> | 2017 | Ghana | 2017 | RDS | Identified as a woman, biologically male, and have had anal or oral sex with another man in the last 12 months | 28.1% (9/31) | 17.4% (650/3723) |
| **Mozambique MoH**<sup>51</sup> | 2018 | Mozambique | 2017 | PLACE | Identified as a woman or transgender, assigned male at birth | 12.7% (15/118) | 9.9% (51/517) |
| **Poteat et al.**<sup>12</sup> | 2017 | Burkina Faso | 2013 | RDS | Self-identified as transgender or female/woman, and assigned male sex at birth | 2.8% (3/108) | 3.3% (18/552) |
|  |  | Côte d'Ivoire | 2016 |  |  | 25.5% (76/298) | 8.0% (61/766) |
|  |  | Gambia | 2011 |  |  | 50.0% (2/4) | 9.0% (18/200) |
|  |  | Lesotho | 2014 |  |  | 59.2% (42/71) | 28.6% (130/455) |
|  |  | Malawi | 2012 |  |  | 16.0% (12/75) | 15.2% (40/263) |
|  |  | Senegal | 2015 |  |  | 37.2% (74/199) | 27.8% (147/528) |
|  |  | Eswatini | 2011 |  |  | 14.2% (17/120) | 18.6% (38/204) |
|  |  | Togo | 2013 |  |  | 17.6% (9/51) | 8.5% (53/626) |
| **Angola MoH**<sup>53</sup> | 2018 | Angola | 2017 | PLACE | Identified as a woman, and assigned male at birth | 9.0% (8/89) | 2.0% (20/1016) |
| **Hakim et al.** <sup>52</sup> | 2018 | Mali | 2015 | RDS | Identified as a woman, assigned male sex at birth, and reported having had insertive or receptive anal sex with a man in the past 6 months | 24.3% (9/37) | 9.7% (12/124) |
| **Jobson et al.** <sup>59</sup> | 2018 | South Africa | 2017 | Snowball | Self-identified as female or transgender, biologically male, and as having sex with men | 57.1% (12/21) | 31.3% (47/150) |
| **Sandfort et al.** <sup>60</sup> | 2019 | Kenya | 2016 | Convenience | Did not identify as male, assigned male sex at birth, and ever had sex with a man | 25.0% (8/32) | 14.6% (20/137) |
|  |  | Malawi | 2016 |  |  | 31.6% (12/38) | 17.9% (15/84) |
|  |  | South Africa | 2016 |  |  | 57.6% (38/66) | 36.9% (90/244) |
| **Liberia MoH** <sup>63</sup> | 2019 | Liberia | 2018 | RDS | No inclusion criterion provided | 27.6% (80/290) | 37.9% (240/633) |
| **Fearon et al.** <sup>13</sup> | 2020 | South Africa | 2017 | RDS | Assigned male at birth and current identity was female or transgender. Sex with a man in last 12 months | 44.6% (20/44) | 38.6% (90/233) |
| **Malawi MoH** <sup>25</sup> | 2020 | Malawi | 2020 | RDS | Self-identified as female or transgender. Engaged in sexual activities with other men in the past 12 months | 13.5% (73/540) | 12.5% (178/1425) |
| **Nigeria MoH** <sup>49</sup> | 2020 | Nigeria | 2020 | Venue-based | Identifies him/herself as a transsexual and undertakes sexual activity with a man | 18.5% (775/4190) | 21.1% (926/4397) |
| **Rwema et al.** <sup>54</sup> | 2020 | Rwanda | 2018 | RDS | Self-identified as transgender or a woman, assigned male sex at birth, and reported having had insertive or receptive anal sex with a man in the past 12 months | 9.4% (10/106) | 10.2% (64/630) |
| **eSwatini MoH** <sup>48</sup> | 2021 | Eswatini | 2020 | RDS | Identified as female. Report have insertive or receptive anal sex with a man in the past 12 months | 41.2% (7/17) | 20.2% (84/415) |
| **Smith et al.** <sup>55</sup> | 2021 | Kenya | 2017 | RDS | Male sex assignment at birth, self-identify as female or transgender, and having had consensual anal or oral sexual activity with a man in the previous 12 months | 40.0% (28/70) | 28.9% (151/522) |
| **Sullivan et al.** <sup>56</sup> | 2021 | South Africa | 2016 | Convenience | Male sex at birth and current gender identity as female or transgender, and report having had insertive or receptive anal sex with a man in the past 12 months | 59.1% (13/22) | 41.8% (110/263) |
| **Harris et al.** <sup>57</sup> | 2022 | Zimbabwe | 2019 | RDS | Born biologically male; self-identified as a woman, transgender, or genderqueer, and engaged in anal or oral sex with a man in the past 12 months | 27.5% (92/335) | 21.1% (248/1176) |
| **Sierra Leone MoH** <sup>58</sup> | 2022 | Sierra Leone | 2021 | RDS | A woman who had a gender identity that is different from her sex at birth | 4.7% (23/487) | 4.2% (18/430) |
| **Zambia MoH** <sup>64</sup> | 2023 | Zambia | 2021 | RDS | Self-identified as a woman, biologically male at birth and self-reported anal or oral sex with biological male in past 6 months | 17% (77/453) | 13.5% (147/1088) |

RDS=respondent driven sampling <sup>38</sup>; PLACE=Priorities for Local AIDS Control Efforts <sup>39</sup>

**Supplementary Table S2:** Estimates of HIV prevalence among transgender men (TGM) compared with transgender women (TGW), cis-gendered men who have sex with men (cis-MSM), and 15-49 total population. In each row, estimates in bold are from the same study indicated in the ‘Study’ column. Transgender women and cis-MSM estimates in italics are the best available comparator estimates from other studies. Total population HIV prevalence estimates are from UNAIDS estimates derived using Spectrum and Naomi models.

| Study | Area | Year | HIV prevalence (%) |  |  |  |  |
| --- | --- | --- | --- | --- | --- | --- | --- |
|  |  |  | TGM | TGW | cis-MSM | 15-49 women | 15-49 men |
| Benin MoH <sup>36</sup> | Benin | 2020 | **9.5** | **26.4** | *7* <sup>49</sup> | *1.1* | *0.5* |
| Mujugira et al. <sup>42</sup> | Kampala, Uganda | 2020 | **4.1** | *20.0* <sup>17</sup> | *12.7* <sup>63</sup> | *7.4* | *4.8* |
| Botswana MoH <sup>27</sup> | Botswana | 2017 | **6.8** | **45.5** | **14.9** | 26.2 | 16 |
| Malawi MoH <sup>43</sup> | Malawi | 2021 | **23.9** | **62.3** | *12.5* <sup>26</sup> | *9.3* | *5.6* |
| Sierra Leone <sup>40</sup> | Sierra Leone | 2021 | **1.11** | **4.36** | **3.4** | *1.8* | *1* |

## Notes

### Competing Interest Statement

The authors have declared no competing interest.

### Author Declarations

The Research and Ethics Committee of Imperial College London gave ethical approval for this work (#6473559)

### Summary of Updates

Updated for submission version to AIDS

